# Doctors Experiences of Interruptions in the Digital Age and the Impact on Clinical Care in England

**DOI:** 10.64898/2026.10.26.26366492

**Authors:** Chase Staras, Antonia Rich, Ann Griffin, Shaun Boustani

## Abstract

**Objective:** Interruptions are frequent in clinical care globally and associated with inefficiency, cognitive burden, and patient safety risks. Existing research into interruptions reflects paper-based workflows, and “digital distraction” studies often focus on personal device use. There is little research regarding interruptions produced by mandated digital infrastructures such as Electronic Health Records (EHRs) and clinical messaging platforms. This study aims to examine how mandated digital systems shape the form, experience, and meaning of interruptions in English outpatient care, and how doctors attempt to manage these demands.

**Methods and Analysis:** We conducted a qualitative analysis of focus group and interview data from a larger study of interruptions in outpatient care. Participants were 42 UK outpatient care doctors across specialties and experience levels, recruited via a screener survey and stratified into six groups based on experience level and self-reported patient complexity (trainee, 0-19 years, and 20+ years’ experience, each divided into high and low complexity). Data were analysed using Reflexive Thematic Analysis, with interpretation informed by the combined lens of Cognitive Load Theory and Social Identity theory; a pairing not previously applied together to interruptions, capturing both the cognitive and social-relational dimensions of digital interruptions. Analysis as structured deductively around these theories while remaining open to inductive, novel insights.

**Results:** Four themes captured digitally mediated interruption. Mandated systems normalised constant availability, making interruption a persistent condition of work. Digital tools acted as cognitive amplifiers, increasing extraneous cognitive load through fragmented interfaces, multi-window navigation, and technical unreliability, with perceived safety risks around patient-context switching. Digital coordination occurred without social cues, weakening visibility and contextual signals that support judgements of relevance, urgency, and responsibility, contributing to interpersonal friction. Doctors sought to reclaim control through individual strategies, including limiting exposure to channels and selectively embracing digital prompts where they supported safety or care quality.

**Conclusion:** Mandated digital infrastructures transform interruption into continuous, cognitively disruptive, and socially ambiguous demands. Mitigation requires system- and organisation-level solutions beyond individual workarounds.

**Key Messages:** *What is already known on this topic:* Interruptions are a known source of cognitive burden and safety risk in clinical care, but research has focused on paper-based workflows and personal device distraction, leaving interruptions from mandated systems like EHRs poorly understood.

*What this study adds:* This study shows mandated digital systems turn interruption into a constant background condition of work, increasing cognitive load through fragmented interfaces and weakening the social cues doctors rely on to judge urgency and responsibility.

*How this study might affect research, practice or policy:* As healthcare becomes increasingly digitally mediated, existing boundaries around availability, communication etiquette, and patient safety need to be revisited as an organisational priority, rather than a challenge for individual doctors to manage alone. This includes clearer digital communication norms and boundaries, safer alert and task-resumption design to reduce patient-context switching risk, and policies that formally protect time for cognitively demanding digital work.

## Introduction

Interruptions are a pervasive feature of clinical practice, defined as a break in the performance of a task, initiated by an external stimulus, that temporarily diverts attention from the primary activity before it is resumed (1). Interruptions are recognised globally as a source of inefficiency and mental burden for healthcare workers, and increases patient safety risks. Interruptions may originate from multiple sources, including other healthcare professionals, patients and their families, alarms and technology, or administrative demands (2,3). Empirical research across hospital settings consistently demonstrates that staff are frequently interrupted during core clinical activities, including patient care, medication administration, laboratory tasks, and surgery (4). Such interruptions have been documented across a range of environments, including critical care (5), inpatient wards (6), and medication administration contexts (7), suggesting that an interruption is not incidental but embedded within the everyday coordination of clinical work (8). Importantly, frequent interruptions have been associated with increased medication errors (9), divided attentional resources (8,10), and delayed task completion (4,11), positioning interruptions as a persistent and hazardous feature of contemporary healthcare delivery.

However, most existing interruptions literature has developed within clinical contexts organised around paper-based systems and relatively stable models of work (Rivera & Karsh, 2010; Teigne et al., 2023). In these accounts, interruptions are typically conceptualised as discrete external events (e.g., a phone ringing, a colleague approaching, an alarm sounding) that momentarily disrupt a bounded primary task, such as a medication round or documentation episode (6,12). This framing assumes neatly divided clinical work in which a primary task can be clearly identified, temporarily paused, and then resumed. Yet these insights rest on assumptions about workflow that no longer reflect the fluid, digitally mediated nature of contemporary clinical practice

Over the past two decades, healthcare systems have undergone large-scale digital transformation, with Electronic Health Records (EHRs) now central to documentation, communication, and care coordination globally (13,14). Clinical work is increasingly conducted through digitally mediated systems that structure how tasks are initiated, prioritised, paused, and resumed (15). As a result, interruptions no longer occur solely as physical interpersonal or environmental disruptions, but also through the routine operation of digital infrastructures.

These digitally mediated interruptions differ in important ways from traditional, face-to-face disruptions. From a cognitive perspective, interruptions embedded within digital systems often occur alongside complex interface navigation, fragmented documentation, and ongoing task switching, all compounding attentional demands and making task resumption more effortful (16,17). From a social perspective, digital interruptions frequently lack the contextual cues that accompany in-person interactions, obscuring intent, urgency, and relational context (18,19). The persistence and visibility of digital communication channels also shape emerging norms around responsiveness, subtly reinforcing expectations of constant availability as part of professional identity (20,21). In this way, digital systems not only increase the frequency of interruptions; they also change how interruptions are cognitively processed and socially interpreted.

Early empirical evidence supports this shift. Studies have demonstrated that the introduction of EHRs is associated with increased task switching and workflow fragmentation, altering both the frequency and form of interruptions experienced by healthcare professionals globally (15,18,22). Similarly, the expansion of instant messaging and alert-based communication systems has introduced persistent channels of contact, enabling interruptions to occur asynchronously, repeatedly, and across spatial boundaries (23,24).

Despite this emerging evidence, most literature addressing “digital distraction” in healthcare has focused on personal device use, such as smartphones accessed for non-clinical purposes during shifts (25–27). This body of work often frames disruption in terms of professionalism, boundary management, or inappropriate technology use. This emphasis has had the unintended effect of obscuring a distinct and increasingly salient phenomenon: interruptions generated through mandated digital systems that are integral to clinical care.

Mandated technologies, such as EHRs and clinical messaging platforms, are organisationally sanctioned, embedded within formal care processes, and are needed for documentation, communication, and decision-making (28). As such, interruptions arising from these systems cannot be readily understood as optional, avoidable, or individual in nature (8). Yet comparatively little attention has been paid to how these mandated digital infrastructures shape interruption patterns, cognitive demands, and doctors’ everyday experiences of work.

To address this gap, the present study offers an original qualitative examination of digitally mediated interruptions in outpatient care across England, with particular attention to those generated through mandated systems such as EHRs and clinical messaging platforms. At their core, these interruptions are cognitive events, imposing attentional and processing demands that disrupt task performance. These processes are well documented in Cognitive Load Theory (CLT) (29,30), which holds that working memory (the limited mental system that temporarily holds and manipulates information during active tasks) has a finite capacity, and that demands placed on it beyond the primary task risk degrading performance. The study draws on CLT to examine how system complexity, fragmented workflows, and frequent tasks switching shape doctors’ attentional demands. However, these cognitive effects do not occur in isolation. Whether, when, and how an interruption is attended to is shaped by the social environment in which it occurs, including expectations placed on doctors to be reachable, responsive, and accountable within their professional role (8). To capture this, the study also draws on Social Identity Theory (SIT) (31), which holds that individuals derive part of their sense of self from the groups and roles they belong to, and regulate their behaviour to match the norms and expectations attached to those roles. SIT is used here to examine how digitally mediated interruptions contribute to evolving norms of responsiveness and availability that become embedded within contemporary clinical identities. This study was guided by the following research question:

In what ways do mandated digital systems shape the form, experience, and meaning of interruptions in everyday outpatient care settings, and how can they be managed?

## Methods

### Design

This study reports a qualitative secondary analysis of data generated as part of a larger study examining interruptions in outpatient clinical work, involving focus groups and interviews with 42 outpatient doctors in England. The parent study was analysed using Framework Analysis, with broader findings reported in our companion papers (8,32). The present analysis focuses specifically on digitally mediated interruptions arising from EHRs, instant messaging systems, electronic task lists, and other mandated digital platforms. Ethical approval was obtained from UCL Research Ethics Committee (ID: 1132).

### Participants and Recruitment

A screener survey was distributed via a large medical defence organisation, capturing demographic and professional characteristics, including experience, perceived job complexity, interruption frequency, and interruption management strategies (33). Participants who expressed interest in follow-up qualitative participation provided contact details and consent for re-contact. Participants volunteered with no prior relationship to the research team, and were informed only that the study concerned interruptions and their management.

Survey data were used to stratify participants into six groups by experience level (trainee; 0–19 years post-qualification; 20+ years post-qualification) and self-reported patient profile complexity (high; low), producing a 3 × 2 stratification matrix (see Table 1). The thresholds used to define the experience groups were based on existing research identifying the point at which experience begins to influence clinical performance. This purposive stratification, rather than convenience sampling, was informed by prior evidence that experience and complexity both shape the cognitive impact of interruptions (34,35), and supported a heterogenous sample across seniority and case-mix.

**Table 1.** Participant numbers, grouped by experience level and self-reported patient profile complexity.

| Group | Experience | Patient Complexity | N<sup>Participated</sup> | Sessions |
| --- | --- | --- | --- | --- |
| 1 | Trainee | High | 3 | 3 Interviews |
| 2 |  | Low | 2 | 2 Interviews |
| 3 | 0-19 Years | High | 10 | 3 Focus Groups |
| 4 |  | Low | 5 | 1 Interview; 2 Focus Groups |
| 5 | 20+ Years | High | 13 | 1 Interview; 3 Focus Groups |
| 6 |  | Low | 9 | 1 Interview; 3 Focus Groups |

**Table 2.**
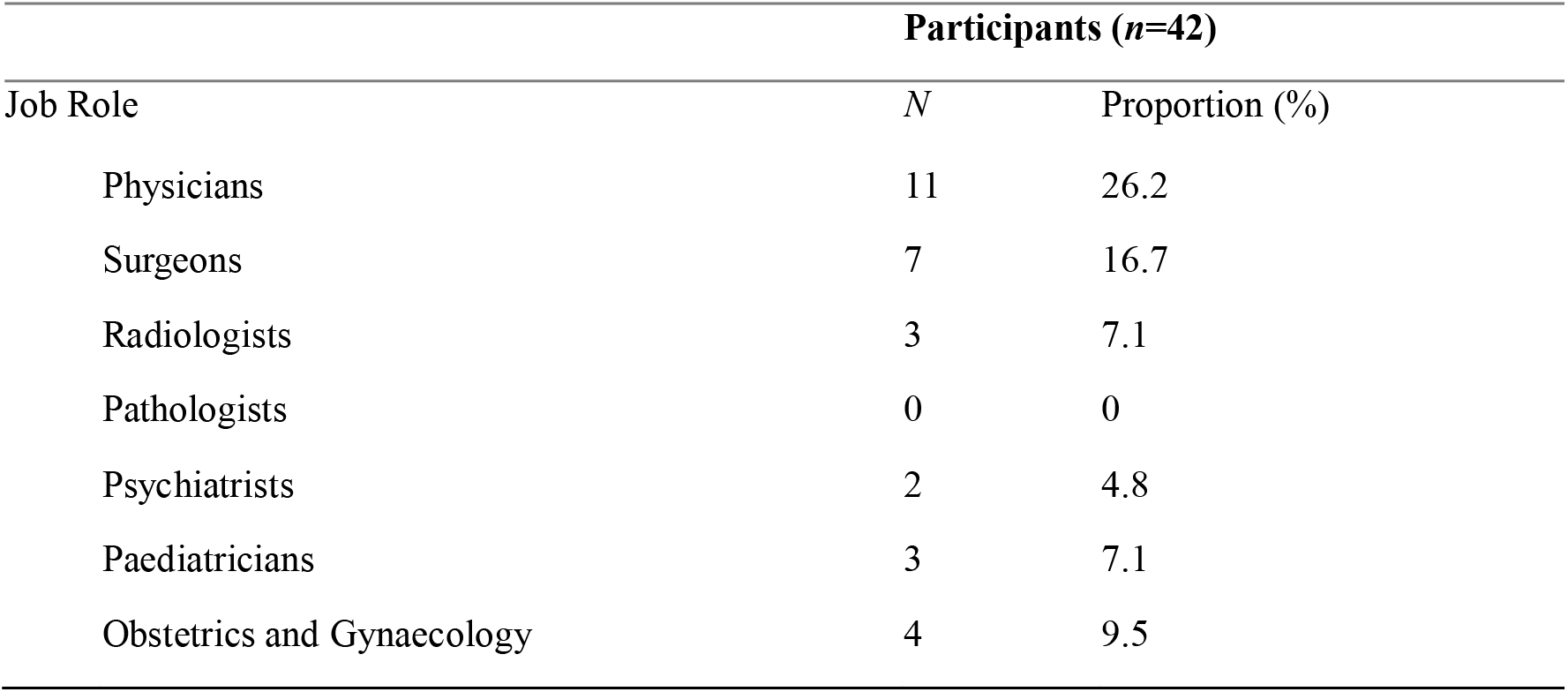
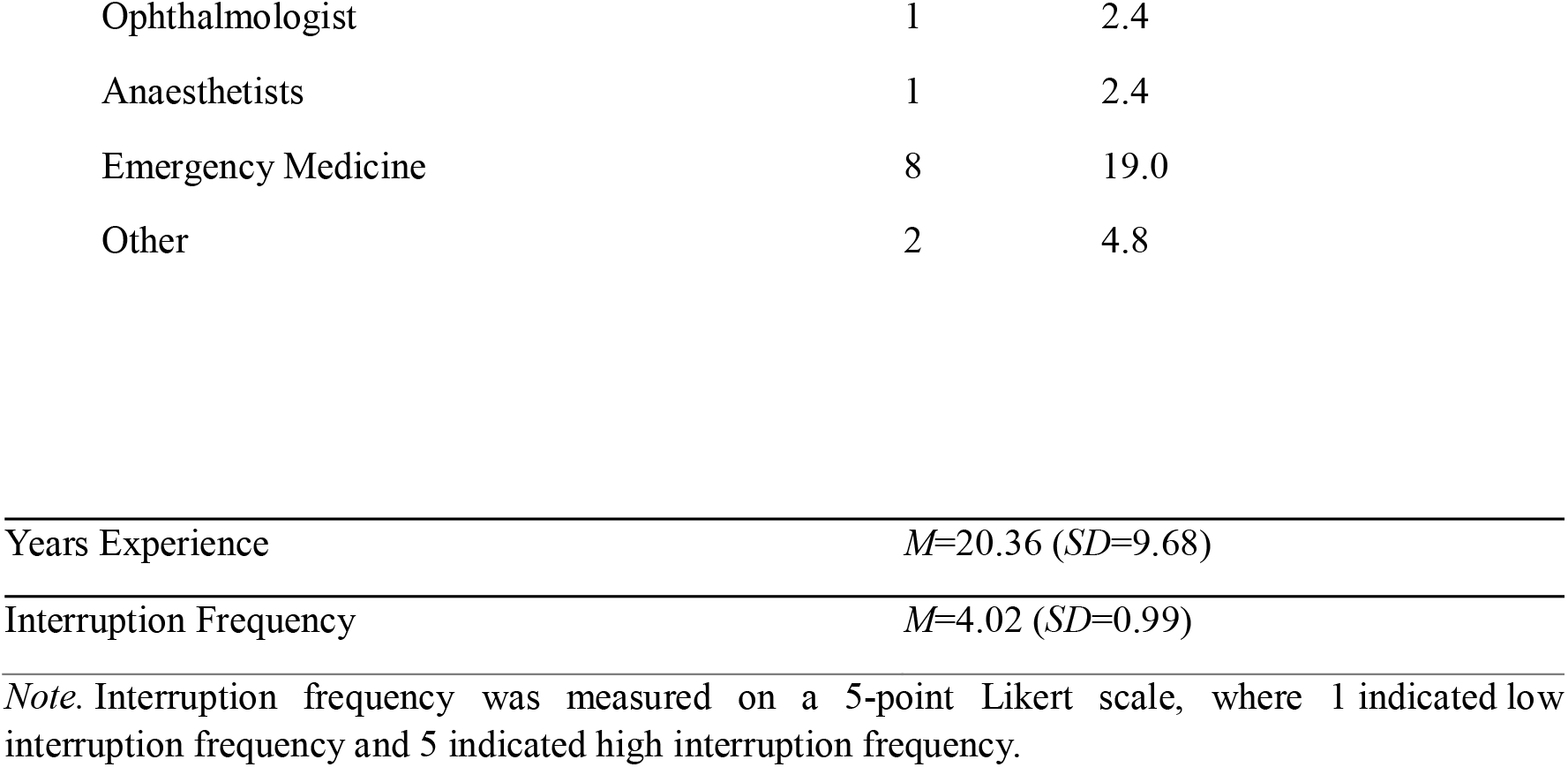
Demographic and professional characteristics of recruited participants.

| Job Role | Participants (n=42) |  |
| --- | --- | --- |
|  | N | Proportion (%) |
| Physicians | 11 | 26.2 |
| Surgeons | 7 | 16.7 |
| Radiologists | 3 | 7.1 |
| Pathologists | 0 | 0 |
| Psychiatrists | 2 | 4.8 |
| Paediatricians | 3 | 7.1 |
| Obstetrics and Gynaecology | 4 | 9.5 |
| Ophthalmologist | 1 | 2.4 |
| Anaesthetists | 1 | 2.4 |
| Emergency Medicine | 8 | 19.0 |
| Other | 2 | 4.8 |

|  |  |
| --- | --- |
| Years Experience | $M=20.36$ ($SD=9.68$) |
| Interruption Frequency | $M=4.02$ ($SD=0.99$) |

*Note.* Interruption frequency was measured on a 5-point Likert scale, where 1 indicated low interruption frequency and 5 indicated high interruption frequency.

Participants within each stratum attended group-specific focus groups or, where scheduling did not permit this, individual interviews. In total, 42 participants were recruited across the six strata, contributing to 19 data collection sessions (see Tables 1 & 2). Recruitment concluded when the sample size was judged to provide sufficient information power for the study’s aims (36).

### Data Collection

Data were collected through focus groups (2-5 participants) and individual semi-structured interviews (approx. 55 minutes) conducted online via Microsoft Teams in 2025, and facilitated by SB (Cognitive Psychologist) and CS (Social Psychologist). The topic guide explored types and sources of interruptions, cognitive impacts, social dynamics, and strategies (see Appendix 1). Transcripts were auto-generated using Microsoft Teams and checked manually against audio recordings. Brief notes were made to support later analysis.

### Data Analysis

Data relating specifically to digital interruptions (24 participants) were re-analysed using Reflexive Thematic Analysis (RTA) (37) in NVivo 14 by CS. RTA was chosen for its capacity for more interpretive, theoretically targeted engagement with this focused subset. Theme development combined inductive analysis with theoretical interpretation informed by CLT and SIT, examining both attentional processes and the social meanings attached to digital interruptions. Findings and draft recommendations were shared with a subset of participants to assess resonance with clinical experience and ensure practical relevance, consistent with qualitative best practice (38).

### Reflexivity

SB’s (male) background in cognitive psychology and medical education informed attention to task complexity and the cognitive consequences of interruptions. CS’s (male) background in social psychology informed analysis of interpersonal norms and professional identity. Ongoing team discussion supported reflexive interrogation of cognitive and social perspectives throughout analysis.

## Results

Reflexive Thematic Analysis produced four themes (see Table 3 for an overview). Each theme is discussed in turn below. All names reported are pseudonyms.

**Table 3.**
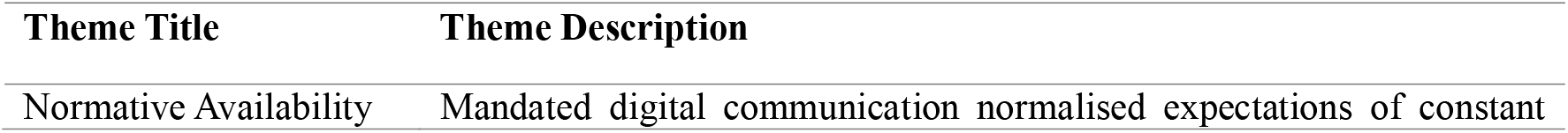
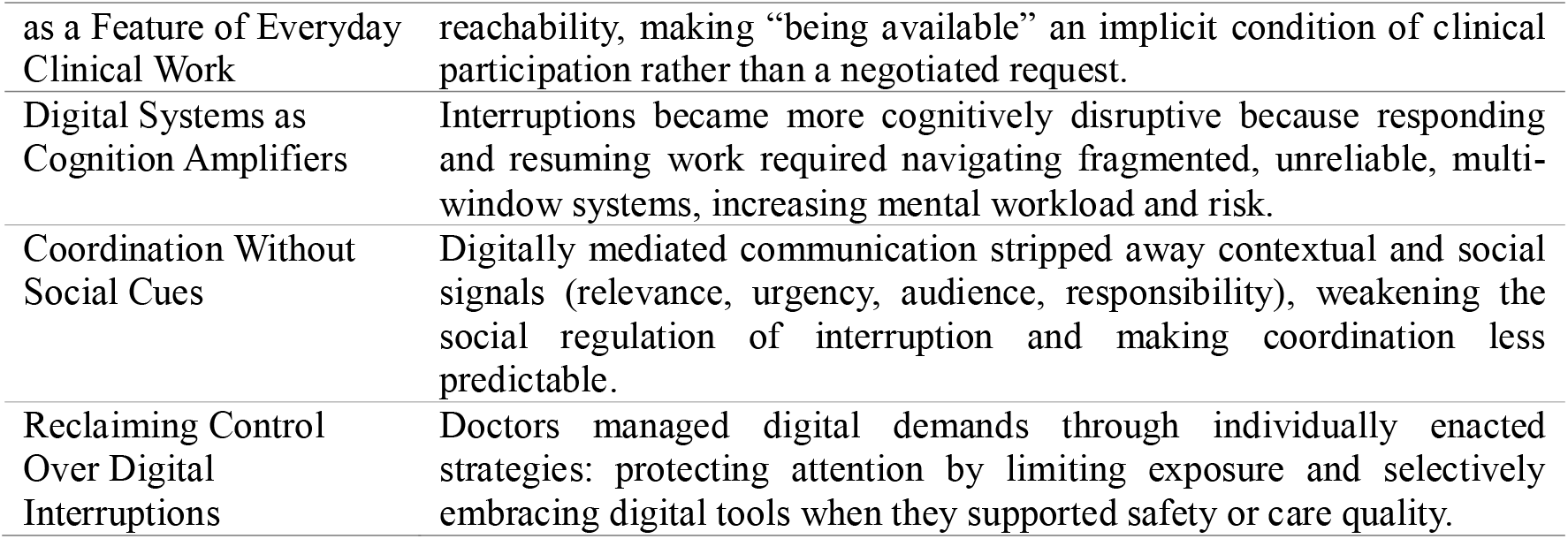
Results of the Reflexive Thematic Analysis.

### Theme 1: Normative Availability as a Feature of Everyday Clinical Work

Participants described working in environments where availability was widely assumed as a routine feature of everyday clinical practice. Rather than being limited to specific roles or moments, digital communication created an expectation that doctors could be contacted at any point during the working day. Availability was therefore experienced not as something negotiated or requested, but as an implicit condition of clinical work:

> “*People see that you are logged onto a computer and then automatically assume that you are available to speak to them, e-mail them back*” (Eimear, Stroke Consultant, Group 3).

In this account, availability is not explicitly demanded but inferred through system presence, with digital visibility overriding contextual cues about workload or competing responsibilities.

This expectation of reachability was reinforced by multiple communication platforms operating in parallel - phones, bleeps, electronic messaging systems, and email - often without shared guidance about which channel should be used in which circumstances. Owen captured the practical burden of this, and the limited control doctors had over communication norms:

> “*It’s multiple channels open, isn’t it? You’re getting WhatsApp emails, physical interruptions, telephone calls, maybe some in-hospital systems that ping up your work list […] The problem with that is somebody will decide there’s some other channel that we will have to use and then we’ll be forced to adapt to it*” (Owen, Clinical Oncologist, Group 6).

The absence of agreed communication norms redistributed the problem of triage onto individual doctors who could no longer rely on any single channel as the place where urgent information would surface. This left many feeling compelled to monitor ever channel simultaneously for fear of missing something important:

> “*Sometimes you think, Oh my God, if I don’t have all 7 methods of communication open at once. I’m going to miss something because actually somebody could contact me about something quite important and urgent on any one of these devices*” (Catrina, Paediatric Oncology Consultant, Group 3).

It is through these everyday practices that mandated digital systems help produce a pervasive culture of immediacy, in which availability is normalised and difficult to refuse, even in the absence of explicit organisational expectations.

### Theme 2: Digital Systems as Cognition Amplifiers

Within this context of constant incoming demands, participants described a marked increase in the cognitive effort required to navigate mandated digital systems. Thus, while interruptions were disruptive in their own right, participants emphasised that digital systems amplified the cognitive workload involved in managing them:

> “*So what was previously either piece of paper or few clicks on the computer? Now it takes much longer and I’ve many more windows for single patient […] Not allowing me to focus on one thing […] and then if I’ve opened a window because I was listing the previous patients for operation, and now I’m not listing. I need X-ray. Then it means closing this window, open another window or a swap in between windows and then I tend to find myself a little bit lost. Have I done the letter or not? Have I booked in for this MRI or not? And this is this is mentally exhausting*” (Baxter, Oncology Consultant, Group 6).Several participants contrasted this with paper-based work, where information could be kept visible and interruptions managed without dismantling the entire workspace. In digital systems, however, rather than simply pausing one task to respond to another, doctors described having to close down multiple screens, switch between systems, and later actively reconstruct where they had been in their original task, placing additional demands on attention and working memory. This combination of interruption and system navigation introduced new risks around task resumption:
>
> “*It just takes your flow and it makes you feel undervalued because you shouldn’t have some of these bureaucratic or administrative problems to deal with. And there are some specific risks, you know. So if you’ve got six IT systems and you’ve put Mr X on all of them ready for the clinic or the consultation, but then the radiographer comes in to ask about Mr Y and you quickly look him up in one of the systems […] 20 minutes later when you’re back talking to Mr Jones, do you miss that you’ve got the wrong one on the screen*” (Owen, Clinical Oncologist, Group 6).

In this way, interruptions created a dual cognitive burden. Doctors had to manage the clinical problem and the additional work of navigating complex digital environments. This dual complexity made task-switching more effortful and recovery more uncertain than in paper-based work.

### Theme 3: Coordination Without Social Cues

Participants described how digitally mediated communication reshaped the social coordination of interruptions by stripping away social context. This weakened cues for judging urgency, relevance, and appropriateness that had previously helped regulate interruptions:

> “I suppose you can’t really tell until you physically get your phone out and look at it or you click on the relevant secure chat message […] *We have a secure chat function […] you’ll probably find that the username you’ve attached to that specialty has got 10 or 15 users inside that. So what becomes a very simple question can very quickly become a whole stream of responses […] and you don’t really know, at that point, who it’s for or who’s meant to pick it up*” (Mason, Emergency and Acute Medicine Consultant, Group 4).

This ambiguity was compounded by multi-user chats that blurred intent and responsibility, often making it harder to judge who a message was directed at or how urgently it should be treated. Participants also described how digitally mediated work removed visible social cues that had previously signalled clinically significant, non-interruptible activity:

> “*We do basically a virtual clinic where we’ll go through sort of 30–40 patients in the morning. To other members of the team we just look like we’re sitting in front of a laptop. So you’re ripe to be interrupted, but actually you are engrossed in decision making and you just don’t happen to have a patient in front of you. And having the cue of a patient in front of me is easier to get back to, than when I’m on a computer and then interrupted*” (Megan, Urology Consultant, Group 6).

Without the social protection afforded by a visibly present patient, digitally mediated clinical work became easier to misrecognise as interruptible, weakening the social pressure that might otherwise restrain interruptions. These accounts suggest that digitally mediated communication reshapes coordination by eroding the cues that previously helped regulate interruption and responsibility.

### Theme 4: Reclaiming Control Over Digital Interruptions

Participants described two main strategies for managing digital demands: 1) limiting exposure to digital channels; and 2) selectively embracing the benefits of digital tools. These strategies were largely individual and situational rather than formally supported by organisational systems.

For many participants, the most immediate response was to limit exposure by silencing devices, closing email, or reducing notifications during clinical work. These actions were framed not as disengagement, but as necessary measures to preserve focus. Some also made their availability more legible to others through informal signals:

> “*My current secretary’s former colleague […] he used to sit at his desk, and if he was in a mood to take a question he would be sitting there normally. But if his tie was draped over his left shoulder, that was his sign for saying do not talk, walk away […] some people do put on their e-mail signatures ‘this is how to get me, this is my timetable’ […] I quite often shut my emails when I’m seeing patients in clinic*” (Owen, Oncologist, Group 6).

These strategies were often fragile, dependent on constant personal vigilance rather than being supported by system design or organisational norms. Alongside boundary-setting, some participants also described a more forward-looking approach, embracing interruptions from digital tools that were perceived to enhance patient care. This included distinguishing interruptions that disrupted workflow from those that introduced safety checks or decision support:

> “*There can be positive digital interruptions as well, so electronic prescribing, there’ll be a pop-up box that’ll come up with: ‘Do you mean to prescribe this? This has already been prescribed? Is this a continuing course?’ Explain your rationale. So there can be safety checks in digital interruptions*” (Darren, Obstetrician & Gynaecologist, Group 4).

Others adopted a more pragmatic stance, acknowledging the disruptive potential of digital devices while also trying to incorporate them into clinical encounters in ways that might improve communication or care:

> “*I suppose it does interrupt things. I say, right, get your phone out. I’m now going to e-mail you. And of course, you tell the parents to get their phone out and they’re going to mention they’re going to answer all their WhatsApp’s and all their other notifications. So yeah, you know, so it is a bit difficult, but I’m also trying to use the technology to improve the way I consult with people. So I think that’s swings and roundabouts*” (Charlotte, Consultant Paediatrician, Group 6).

These accounts suggest that managing digital interruptions did not always mean reducing them. Instead, doctors sought to reclaim control by carving out pockets of protected attention and by reframing digital systems as tools for safer or more effective care.

## Discussion

This study is the first to explore how outpatient doctors experience mandated digital interruptions, applying a combined cognitive and social theoretical framework to specify why these interruptions generate greater burdens than the face-to-face interruptions that characterised former healthcare delivery. This joint CLT-SIT lens offers a transferable framework for researchers examining how technology-mediated demands intersect with professional identity in other high-accountability settings.

### From Episodic to Ambient Interruption

Prior work has largely treated interruption as an episodic disruption, escapable by changing location or context (2,11). Our findings extend this by showing that mandated digital systems create a sustained condition of availability that doctors cannot step outside of, even by physically leaving a ward or working from home. This reframes interruption not as a discrete event, but as a background condition of digitally mediated work; a shift in kind rather than degree from how the phenomenon has previously been conceptualised.

### From Information Overload to Systemic Cognitive Disruption

Our findings align with, and extend, earlier work on workflow fragmentation in EHR-mediated work (15,18,22) in three ways. First, whereas prior research has largely focused on single settings, such as emergency medicine (15) or primary care (22), our findings suggest these dynamics are a general feature of digitally mediated clinical work rather than being confined to particular specialties.

Second, rather than a narrow subset of digital functions such as inbox management, our findings speak to the wider ecology of mandated systems. Interruption risk arises from how multiple systems interact, not from any single tool, positioning digital interruption as a systemic property of contemporary clinical work rather than a problem localised to specific departments.

Third, where existing accounts largely frame the problem in terms of information volume (18), our findings show the burden lies not only in how much information doctors receive, but in the structure of the cognitive work required to manage it: task switching, navigating fragmented interfaces, and verifying that the correct patient is in view. This shift from quantity to structure helps explain why digital interruptions can be effortful and error-prone even when their informational content is simple, and links this cognitive burden directly to safety risk, particularly around patient-context switching.

### From Communication Breakdowns to the Erosion of Social Regulation

Existing literature has noted fragmentation in digital healthcare communication (15,18,22) and coordination challenges in digital channels more broadly (19,21), but has not directly theorised how the social norms and visible cues that once regulated interruption are altered when work is digitally mediated. Our findings address this gap. In face-to-face settings, interruption is regulated through visibility, timing, and shared situational awareness (5,11), whereas digital interruptions frequently arrive stripped of these signals, with multi-user chats further obscuring who is being addressed. From a SIT perspective, this matters because professional norms of responsiveness are enacted through everyday interactional cues (31). When these are absent, doctors are left to infer expectations and manage accountability individually. This illuminates a novel avenue through which digitally mediated interruption disrupts clinical workflows: not only as a problem of information overload, but of social regulation.

### From Reduction to Managing Digital Interruption in Practice

Finally, participants also described two broad strategies for managing digital demands: limiting exposure to digital channels, and selectively embracing tools perceived to enhance safety or care quality. Neither strategy reversed the conditions described above. Rather, both represent attempts to make digitally saturated work more tolerable within existing constraints, echoing prior observations that doctors often rely on local workarounds rather than system-level solutions when facing EHR-related burden (15,17). This is consistent with evidence that EHRs often shift interruption management onto doctors rather than embedding protections into workflows (15,18), reinforcing sociotechnical arguments that poorly aligned digital systems externalise coordination and safety work onto users rather than absorbing it into system design (15,18). In practice, doctors experience the steady addition of new digital systems as an accumulation of cognitive load and fragmented attention, layered onto persistent expectations of constant availability, with no organisational imperative to help them manage it (39). This highlights the need for organisational approaches to interruption management.

### Strengths, Limitations, and Future Directions

This study extends interruption research beyond paper-based, task-bounded models (2,11), and narrow foci on personal device distraction (25,26), by examining how mandated digital infrastructures shape interruption across outpatient care. By integrating cognitive (e.g., CLT) and social (e.g., SIT) perspectives, it moves beyond accounts of information overload (18) to show how digital systems transform both the cognitive work of task management (16,17) and the social regulation of attention and responsibility (19,21). The research team’s complementary professional backgrounds, spanning cognitive psychology, clinical practice, health psychology, and social psychology, allowed data to be interrogated through multiple analytic lenses, reducing the risk that findings reflect a single disciplinary standpoint. Finally, emerging findings and draft recommendations were shared with a subset of participants to assess resonance with their clinical experience, a form of member checking that supports the credibility of the analysis (38).

Two limitations should be noted. First, this study draws on qualitative accounts rather than direct observation or system log data. Future research could complement this approach with ethnographic studies or EHR trace data, as used in studies of workflow fragmentation and inbox work (15,22). Second, although the sample spans multiple specialties and experience levels, it is situated within English outpatient care and within particular EHR and messaging infrastructures. However, given that EHR adoption reflects a shared international policy priority rather than a UK-specific phenomenon (40), the mechanisms identified here are likely to extend to other health systems with comparable digital infrastructures, though comparative studies across settings and platforms would help to establish this directly given system configuration and organisational context strongly shape interruption patterns (18,28).

Future research should test intervention strategies at the level of system design and organisational policy, examining how alert hierarchies and task interfaces shape cognitive load (16,17). and how team-level norms might reintroduce shared regulation of interruption (21).

### Implications for Policy and System Design

The findings have important implications for digital health policy and EHR design. Current approaches often frame interruption as an unavoidable consequence of connectivity, or as a problem of individual professionalism (25,26). Our findings instead align with sociotechnical critiques showing that EHR-related burden is produced through design choices and organisational arrangements, not merely user behaviour, meaning solutions cannot rest solely on doctors managing their attention better (15,18,28).

There is a need for organisational strategies that reintroduce meaningful boundaries and shared norms for digital communication, echoing calls for system-level approaches to workflow fragmentation (3,17). This includes clearer signalling of urgency and responsibility, better support for task resumption, transparent management of multi-user channels, and protected time for cognitively demanding work (18,20–22). Without such changes, responsibility for coping with digital interruption will continue to fall disproportionately on individual doctors.

### Conclusion

This study shows that mandated digital systems do not merely add new interruptions to existing clinical workflows (2,11). They transform interruption itself: from episodic events into ambient conditions of work, from momentary disruptions into sources of systemic cognitive strain (16,17), and from socially regulated interactions into ambiguous demands stripped of shared cues and norms (19,21). While doctors develop pragmatic strategies to cope with these conditions, these remain individualised and weakly supported by system design (15,18). By conceptualising digital interruption as both a cognitive and social phenomenon, this study advances current debates beyond information overload and personal device distraction. It highlights the need to treat interruption as a design, organisational, and policy problem, not simply an individual one, if digitally mediated healthcare is to support, rather than systematically undermine, attention, coordination, and safety.

## Data Availability

The data are not publicly available as we do not have the necessary permissions for sharing. Anonymised quotations are included in the article.

## Appendices

### Appendix 1: Interview Topic Guide

#### I. Focus Group Interview Guide

*This semi structured guide is intended for online focus groups (grouped by similar levels of expertise to mitigate power dynamics). It is divided into thematic sections to explore the nature of interruptions, their cognitive and social impacts, and strategies used for their management.*

#### Ethics Statement

Hello — I’m XXX, I’ll be leading the interview/focus group today.

Thank you for taking part. Before we begin I need to cover a few important ethical points.

Participation is voluntary and this session will take about 45 minutes. There aren’t any right or wrong answers and we’re just looking to get your opinions and experiences.

You’ve already given electronic consent via the screener; that consent covers your survey responses and the audio-recording for this focus group. If you would like to withdraw for any reason you can do so at any time during the session. Please note, however, that as transcripts will be pseudo-anonymised, it will not be possible to remove contributions made to that point in the analysis.

Recordings and transcripts will be stored securely on UCL systems, and only the research team will have access. We will also be taking notes during the session just to remind us of important information.

If that’s all clear, I’ll start the recording. Do you have any questions or objections to being recorded?

#### A. Introduction (5 minutes)

- “As you know, we are interested in how unexpected interruptions—such as a colleague asking a quick question or a phone call in the middle of a consultation—affect your work. We also want to understand how these events might change the way you feel mentally, how they might influence interactions among team members, and how you manage an interruption once its occurred.”

#### B. Experiences with Interruptions (15 minutes)

##### Nature of Interruptions

- “To start, could you describe a typical interruption in your clinic. How often do these occur?”
- “What kinds of interruptions do you encounter most frequently (e.g., trainee queries, phone calls, unexpected staff or patient interactions)?”

##### Context & Impact

- “Are there any particular conditions (time of day, workload, clinic structure, staffing level) that make interruptions more or less easy to cope with?
- “How do you think these interruptions influence your work

- and what about how they influence patient care and safety?”
- “What do you think makes an interruption particularly disruptive?

- Is it the context, timing, who it comes from, or something else?

#### C. Cognitive Impacts and Task Demands (15 minutes)

- **Cognitive Load: I’d now like to ask some questions regarding cognitive or mental impacts of interruptions.**
- **Explaining the Concept:**

- “Sometimes, we use the term ‘cognitive effort or cognitive load’ to describe how much thinking or concentration is needed to complete a task. We’re interested in how interruptions may change this feeling.”
- “Thinking about a recent experience, did an interruption make it more challenging to get back on track with your consultation, if so, how?”
- “Are there certain parts of your work where interruptions seem to have a bigger impact on your concentration?”
- “Prompt: does the complexity of patient information or the demands of the task influence your experience?”
- **Resumption Strategies:** “What do you do to get back on track after an interruption? Are there any routines or steps you rely on?”

`o` What do you think helps you decide whether to resume immediately, delay, or drop a task after being interrupted?

##### Cog Impact and Mental Wellbeing

“Do you think these demands affect your wellbeing?”

#### D. Social Dynamics and Interpersonal Aspects (15 minutes)

- “We’re also curious about how these interruptions influence your interactions with colleagues. By ‘team communication,’ we mean any conversation or exchange with other staff—before, during, or after a consultation.”
- **Team Communication:**
- “How do interruptions affect your interactions with colleagues?”
- "Do you think interruptions are perceived differently depending on whether they come from someone ‘inside’ your team versus someone from another team or department?"
- If the interrupter is known: “Does your relationship with the interrupter impact the way you manage or respond to an interruption? What about their seniority?”
- **Positive and Negative Impacts:**

- “We are also curious of whether there are any positive effects of interruptions. Have you observed any positive effects (e.g., brief, supportive interactions) when interruptions occur? Under what circumstances?”
- **Prompt: Are there circumstances you don’t mind being interrupted?**
- **Prompt 2: Are there circumstances where you think being interrupted could be helpful?**

- Now looking towards some potentially negative impacts, “Can you share examples when an interruption compromised team cohesion.
- When interruptions go badly, what is usually happening in the background that contributes to this?
- Prompt: When the interruption has occurred, do they sometimes lead to clarifications or, conversely, to misunderstandings?”
- Prompt: Thinking about interruptions, both positive and negative do you feel interruptions sometimes trigger emotions (e.g., frustration, relief, reassurance) that shape how you respond?

#### E. Management and Coping Strategies (10 minutes)

##### Current Practices

Finally, I just wanted to discuss some current strategies, essentially the ways you’ve learned to cope with interruptions once they’ve happened…

- “What strategies have you or your team adopted to manage interruptions effectively?”

- Prompt: In what situations do your strategies for handling interruptions work well, and when do they not?
- How do you know which tasks to prioritise? Can you give an example
- If participants are struggling to prompt on what we know from the survey findings: prevention; cognitive (offloading); social (putting people off).
- “Have you used any structured approaches (e.g., communication protocols like ISBAR) that you believe could work better?”

##### Barriers and Facilitators

- “What barriers prevent the effective management of interruptions in your setting?
- What factors or resources would facilitate better management?”

#### F. Future Directions and Concluding Discussion (5 minutes)

- **Suggestions for Intervention:**

`o` “Based on your experiences, what improvements or interventions would you suggest to reduce the negative impact of interruptions?”
- **Closing Remarks:**

`o` Thank participants for their valuable contributions and reiterate how their feedback will help improve both patient care and team well-being.

